# Analysis of *BRCA1*, *BRCA2* and *PALB2* related Fanconi anemia identifies scope to expand disease phenotypic features and predict breast cancer risk in heterozygotes

**DOI:** 10.1101/2025.05.25.25327887

**Authors:** Sharon E. Johnatty, Emma Tudini, Michael T. Parsons, Kyriaki Michailidou, Maria Zanti, Daffodil Canson, Aimee L. Davidson, Tamar Berger, Rasim Ozgur Rosti, Christian P. Kratz, Reinhard Kalb, Lisa J. McReynolds, Neelam Giri, Marcy Richardson, Tina Pesaran, Jordi Surrallés, Roser Pujol, Babu Rao Vundinti, Merin George, Kara N. Maxwell, Kate Nathanson, Susan Domchek, Moisés Ó. Fiesco-Roa, Sara Frias, Benilde Garcia-de-Teresa, Marjolijn Jongmans, Seema Lalani, Merel Maiburg, Katrina Prescott, Rachel Robinson, Sulekha Rajagopalan, Lot Snijders Blok, Suzanna E. L. Temple, Kathy Tucker, Arleen D. Auerbach, Maria I. Cancio, Jennifer A. Kennedy, Margaret L. MacMillan, Rebecca Tryon, John E. Wagner, Michael Walsh, Nicholas J. Boddicker, Chunling Hu, Jeffrey N. Weitzel, Alexander J. M. Dingemans, Johanna Hadler, Nitsan Rotenberg, Lobna Ramadane-Morchadi, Miguel de la Hoya, Paul James, Thomas Van Overeem Hansen, Maaike P. G. Vreeswijk, Logan C. Walker, Shyam K. Sharan, Douglas F. Easton, Fergus Couch, Agata Smogorzewska, Adam Nelson, Joanne Ngeow, Marc Tischkowitz, Encarnacion Gomez-Garcia, Amanda B. Spurdle, the ENIGMA, BCAC and CARRIERS consortia

## Abstract

Recessive Fanconi anemia (FA) phenotype is used in classification of *BRCA1/FANCS*, *BRCA2/FANCD1* and *PALB2/FANCN* variants with respect to dominant hereditary breast-ovarian cancer syndrome. We assessed its utility by examining the spectrum of phenotypes observed in individuals biallelic for *BRCA1*, *BRCA2* or *PALB2* pathogenic variants, and exploring the relationship between cancer presentation and allele severity score based on variant molecular features.

A data collection instrument comprising 158 Human Phenotype Ontology (HPO) terms was used to document clinical features for individuals with FA from published and/or prospectively collected (total n=172, 43 previously unpublished) phenotypic data. Unique FA-related variants (15 *BRCA1*, 123 *BRCA2*, 22 *PALB2*) were annotated for predicted molecular impact, location, observed splicing or functional impact, and potential in-frame splice rescue. Annotations were used to assign different permutations of allele severity scores, which were assessed for correlation with FA presentation features. The association of *BRCA1* and *BRCA2* allele severity score with magnitude of breast cancer risk in heterozygotes was evaluated using case-control analysis.

Patient-detected features extended beyond the FA ORPHA:84 HPO list, including 84 terms related by hierarchy, and 94 novel terms. Genotype severity score was significantly associated with age at cancer diagnosis in *BRCA2* FA individuals (p=1.8×10^-8^). A similar permutation approach revealed significant differences in magnitude of breast cancer risk according to *BRCA1* and *BRCA2* allele severity score in heterozygotes. Findings indicate potential to redefine the existing list of FA-related HPO terms, and to use an allele severity scoring approach to predict cancer risk in both FA patients and heterozygotes.

## INTRODUCTION

Fanconi anemia (FA) is a rare genetic disease characterized by chromosomal instability due to defective function of proteins involved in DNA repair.^1^ Of the 23 genes implicated in FA, one (*FANCR*/*RAD51*) causes autosomal dominant FA, another (*FANCB*) causes X-linked FA, and the remaining 21 have an autosomal recessive pattern of inheritance where biallelic pathogenic variants cause disease.^2^ In some cohorts, over 90% of FA individuals are associated with recessive inheritance of pathogenic variants in *FANCA*, *FANCC, FANCD2* or *FANCG*. Rarer presentations of FA include biallelic inheritance of pathogenic variants in FA pathway genes known to increase risk of breast and/or ovarian cancer, including *BRCA1/FANCS, BRCA2/FANCD1*, *PALB2/FANCN, RAD51C/FANCO,* and *BRIP1/FANCJ*.^2,3^ The FA pathway is involved in the repair of DNA interstrand crosslink repair, with these breast-ovarian cancer susceptibility genes playing a key role in the homologous DNA recombination-mediated repair of DNA double strand breaks that arise following interstrand crosslink removal.^2,3^

The FA phenotype includes physical abnormalities that variably affect several parts of the body, and increased risk of developing progressive bone marrow failure from the first decade of life.^2,4^ They also have an extraordinarily higher risk of early-onset solid and hematological cancers compared to the general population, as well as increased sensitivity to DNA cross- linking agents such as platinum chemotherapeutics,^2,4^ and to ionizing radiation.^5^

Presence of a variant *in trans* with another known pathogenic variant in the context of a recessive phenotype is used as evidence towards variant pathogenicity, including the classification system of the American College of Medical Genetics and Genomics and Association of Molecular Pathology (ACMG/AMP).^6^ As such, it has been included as an evidence type in the ClinGen Variant Curation Expert Panel (VCEP) specifications for *BRCA1*, *BRCA2* and *PALB2* (https://cspec.genome.network/cspec/ui/svi/). Recommendations for defining FA phenotype included phenotypic features for FA compiled for all FA genes combined as drawn from GeneReviews^®^,^2^ and cancer diagnosis age cut-offs based on literature review for FA individuals bi-allelic for pathogenic variants in these genes specifically.^7^

There is evidence that FA phenotype due to *BRCA1*, *BRCA2* and *PALB2* may be more severe (if not mostly embryonic lethal) compared to that due to pathogenic variants in most other FA genes. Patients with FA due to biallelic *BRCA2* or *PALB2* pathogenic variants have a higher probability of developing cancer, and at much younger ages, compared to other more commonly observed FA subtypes, requiring more intensive surveillance and screening.^8–10^ The cancer spectrum is also unique with embryonal tumors predominating, such as medulloblastoma.^4^ In FA mouse models, biallelic null variants in *Brca1*, *Brca2* and *Palb2* (and also *Rad51C* and *Rad51*) are embryonic lethal (reviewed in Guitton-Sert et al ^11^).

Biallelic *BRCA1* pathogenic variant status was initially considered to be embryonic lethal, but there is emerging evidence that specific combinations of alleles being support viability because they retain sufficient function and/or their deleterious impact is rescued through alternative splicing (reviewed in Hughes et al ^12^). Evidence for prenatal selection against specific combinations of alleles also exists for *BRCA2*. There are no reported individuals biallelic for Ashkenazi Jewish founder *BRCA2* pathogenic variants; previous analysis showed a depletion of FA individuals biallelic for pathogenic variants within the largest *BRCA2* exon 11 that harbors almost half of the reported pathogenic variants in this gene.^4^ There is also evidence to support that some alleles identified in *BRCA2* FA individuals may be hypomorphic in relation to experimentally measured variant impact on splicing or protein function, or may be rescued through alternative splicing.^13–16^

Here, we undertook a study to document the phenotypic and genotypic features of FA individuals with biallelic pathogenic variants in *BRCA1*, *BRCA2* or *PALB2,* to better inform the use of FA phenotype to predict pathogenicity of individual FA-detected alleles in the context of dominantly inherited cancer due to these three genes.

## STUDY DESIGN AND METHODS

The collation and analysis of de-identified data for the overall study was approved by the QIMR Berghofer Human Research Ethics Committee (P1051). Patient data collection performed by external sites for prospective data analysis was approved under the following institutional review board codes: Utrecht University METC 2022-3494 (EG); NCI IBMFS study NCT00027274 (LMcR, BPA & NG); The Rockefeller University AAU-0112 (AA, AS & TB); Spanish Biobank of DNA Repair Syndromes Collection No. C.0002193 (JS & RP); ICMR-National Institute of Immunohaematology NIIH/IEC/23-2019 (BRV & MG); Instituto Nacional de Pediatría INP 053/2020, Institutional Review Board (IRB): IRB00013674 (SF, BG & MÓF-R), Baylor College of Medicine H-41191 (SL); Protocol #376800 (Penn) with HIPAA waiver of consent (KM, KN & SD); Cincinnati Children’s Hospital Medical Center, IRB NCT02143830 (AN); German Cancer Predisposition Syndrome Registry, DRKS00017382 (CPK), Memorial Sloan Kettering Cancer Centre Institutional Review Board (JAK, MIC, MW). All patients included in prospective data collection provided written informed consent for de-identified data collection.

A schematic of the overall study design and workflow is shown in **Figure 1**.

**Figure 1:**
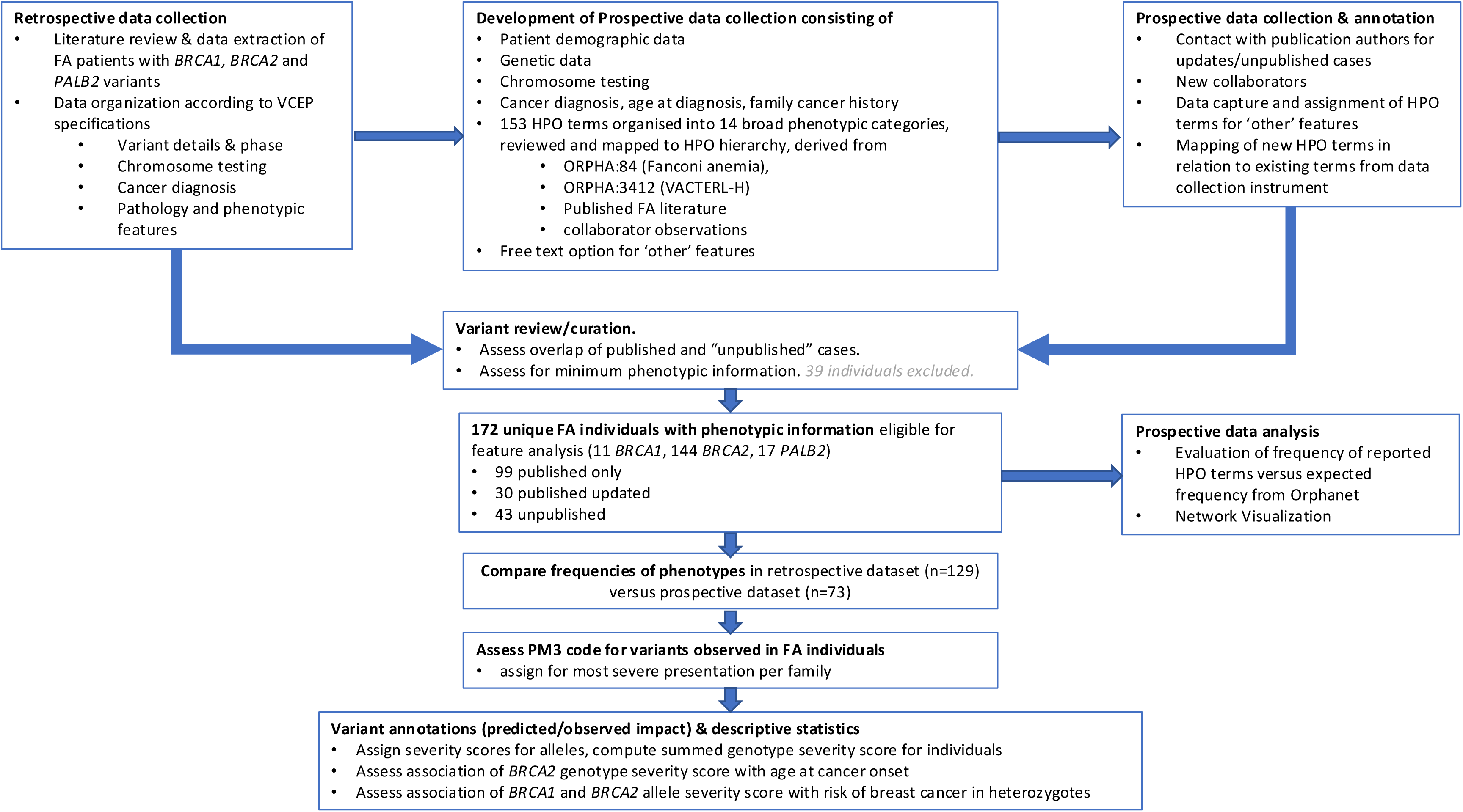
Schematic of overall study design and workflow. See Methods and Supplementary Methods for further details on data collection, annotation, and analysis.

The FA dataset used for descriptive and other analyses comprised two partly overlapping groups: 1) a “retrospective dataset” curated from published reports of FA individuals biallelic for *BRCA1*, *BRCA2* or *PALB2* pathogenic variants; and 2) a “prospective dataset” of FA individuals biallelic for *BRCA1*, *BRCA2* or *PALB2* variants obtained via collaboration that involved systematic clinical data collection for a subset of previously published individuals, as well as for previously unpublished individuals.

### Retrospective data collection

Published reports of individuals diagnosed with FA who harboured *BRCA1*, *BRCA2* or *PALB2* variants were identified using the keywords “Fanconi anemia”/ “Fanconi anaemia” and “FANCS”, “BRCA1”, “FANCD1”, “BRCA2”, “FANCN” or “PALB2” in PubMed.

Retrospective data collection was restricted to reports published from January 2000 to August 2024. References from within these publications were also reviewed for additional information relevant for individual cases. We extracted, where available, patient genotype, sex, age at FA diagnosis, results from chromosomal breakage studies, phenotypic features consistent with FA, cancer diagnoses and outcomes (specifically age of death), pathology findings including toxicities from chemotherapy or radiation treatments, progressive bone marrow failure unrelated to cancer treatment, stem cell transplantation data, family cancer history, and reported sequence variants in genes of interest. We also contacted study authors to clarify variant details if required, and invite them to participate in the prospective data collection arm of the study.

*BRCA1*, *BRCA2* and *PALB2* variant information was corrected to reflect Human Genome Variation Society (HGVS) nomenclature. Variants were first reviewed for evidence of pathogenicity using assertions (and related justifications) in ClinVar records as at January 2024. For variants considered to be of uncertain clinical significance (VUS) after this initial review, or variants absent from ClinVar, evidence for pathogenicity was assessed using ClinGen ENIGMA BRCA1 and BRCA2 VCEP Specifications V1.0 and ClinGen PALB2 VCEP Specifications V 1.1. Following curation for variant pathogenicity, and assessment for any reported FA phenotypic features, we excluded 29 individuals identified from publications (Data_type code PE) and another five patients from publications that remained ineligible after receiving updated information from collaborators (Data_type code PUE). See **Table S1** for justification of exclusions and further details. For another 30 patients identified from the literature, the data from the publication was coded as superseded (Data_type code PS) due to additional updated information provided as part of the detailed prospective collection (Data_type code PU). Lastly, two individuals were identified through subsequent contact with authors to be the same patient, with some resolution of additional characteristics (Data_type annotation PE, overlap).

The retrospective dataset (Data_type codes P and PS) included 129 individuals reported to have FA (10 *BRCA1*, 104 *BRCA2*, and 15 *PALB2*) and: (i) with sufficient details to assign HGVS nomenclature; (ii) biallelic for (likely) pathogenic variants in the same gene, with the exception of two individuals with one (likely) pathogenic variant and a second variant considered after review to be a VUS with high suspicion of pathogenicity (VUS suspicious; see **Table S1** for further information); (iii) with a phenotype broadly consistent with FA diagnosis based on existing definitions that had been documented for use in variant interpretation by the ClinGen ENIGMA BRCA1 and BRCA2 VCEP, drawn from GeneReviews (see Supplemental Information, Supplemental Methods).

### Development of the prospective data collection instrument

Review of the reported features in the retrospective dataset was used to inform development of a comprehensive data collection instrument, that included patient diagnostic and demographic data, response to chemotherapy or radiation therapy, genotype, bone marrow transplantation and clinical phenotype related to FA.

Human Phenotype Ontology (HPO) terms for phenotypes were obtained from the publicly available Orphanet rare disease database (https://www.orpha.net/en/disease), which is closely linked with the HPO project (https://hpo.jax.org/).^17^ HPO annotations, and their expected frequencies within patient groups, defined as ‘very frequent’ (80-99%), ‘frequent’ (30-79%), or ‘occasional’ (5-29%) based on the published literature, were accessed on June 8, 2021 for Fanconi anemia (ORPHA:84), and for the differential diagnosis VACTERL with hydrocephalus (VACTERL-H; ORPHA:3412). A total of 106 HPO terms for FA (ORPHA:84; 13 ‘very frequent’, 10 ‘frequent’ and 83 ‘occasional’) and 33 HPO terms for VACTERL-H (ORPHA:3412; 12 ‘very frequent’, four ‘frequent’ and 17 ‘occasional’), were obtained from Orphanet. Eleven FA phenotypes overlapped with VACTERL-H; six of the 22 unique VACTERL-H terms were classified ‘very frequent’, four were ‘frequent’, and 12 were ‘occasional’. Review of 42 potential additional FA-related terms identified through unpublished observations from collaborators or publications^18–20^ revealed 30 more HPO terms not listed among the ORPHA:84 and ORPHA:3412 Orphanet terms. The resulting 158 HPO terms from all sources and their overlap are shown in **Figure 2**, and **Table S2**.

**Figure 2:**
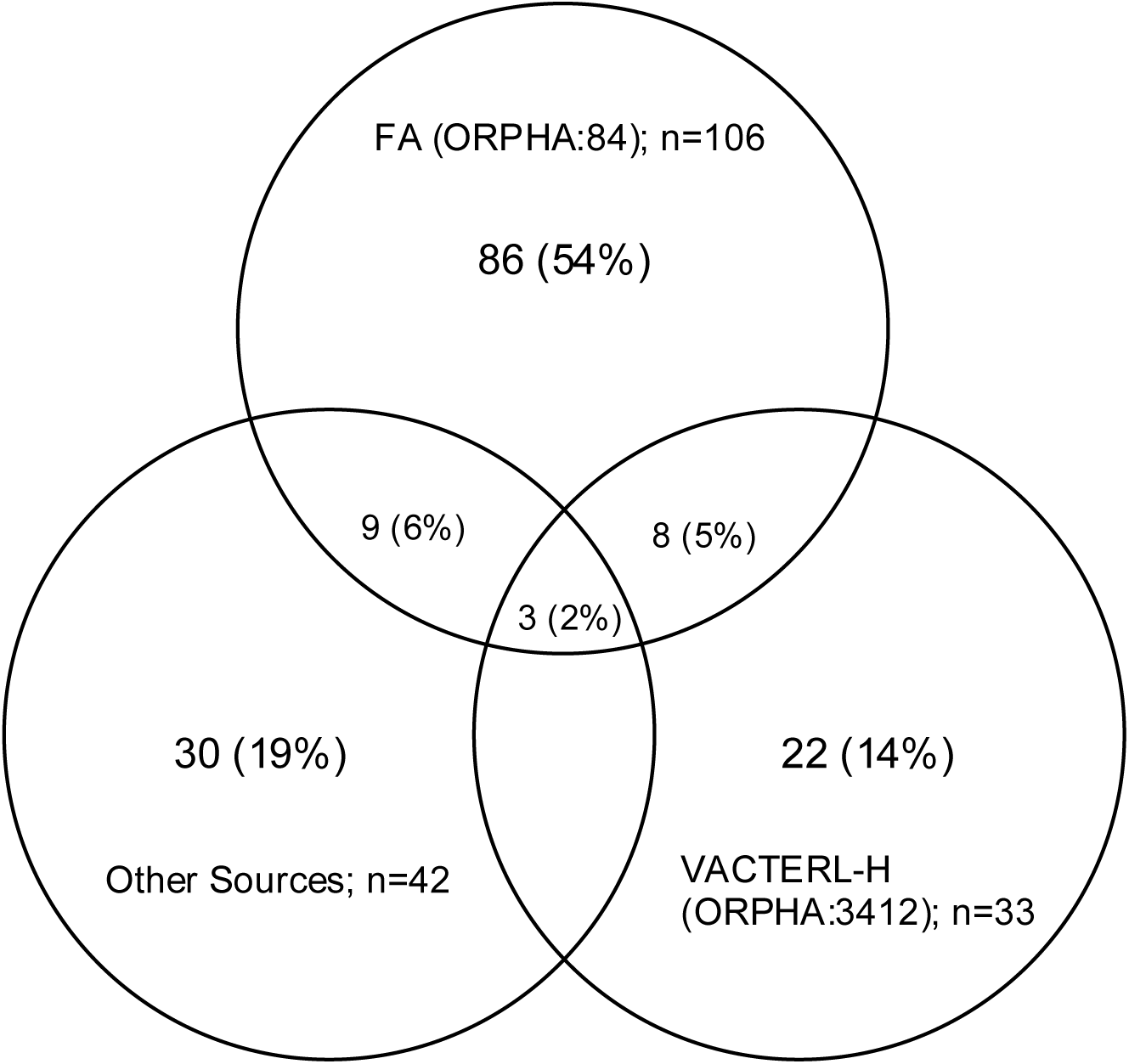
Overlap of 158 Human Phenotype Ontology terms from three major sources. Human Phenotype Ontology (HPO) terms included in the prospective data collection instrument were derived from Orphanet for FA (ORPHA:84, n=106) and VACTERL-H (ORPHA:3412, n=33), and ‘Other sources’ based on a review of published studies and collaborators informing development of the data collection instrument (n=42). Overlap of terms between sources was as follows: 11 VACTERL-H terms overlapped with FA; 12 ‘Other sources’ terms overlapped with FA; 3 terms were common to all three sources. The breakdown of HPO terms according to their sources is shown in Table S2.

Five of the ORPHA:84 terms, HP:0003220 (Abnormality of chromosome stability), HP:0002863 (Myelodysplastic syndrome), HP:0002664 (Neoplasm/Cancer diagnosis), HP:0000478 (Abnormality of the eye), HP:0001871 (Abnormality of blood and blood-forming tissues) are broad phenotypic abnormalities, and were collected in more detail or as discrete phenotypic categories. We reviewed the remaining 153 specific terms for redundancy and mapped them according to the HPO hierarchy for parent-child-grandchild relationships based on HPO releases available as of 5^th^ April 2023. A term was designated ‘parent’ if at least one term on the list was its direct descendent, the latter designated ‘child’, and ‘grandchild’ if a term was a direct descendent of the ‘child’ term **(Table S2)**. We identified 22 parent terms, 41 child terms and three grandchild terms within the list of 153 terms. The finalized list of HPO terms, including annotations for parent-child-grandchild relationships, was collated across fourteen broad phenotypic abnormalities for prospective data collection, shown in **Table 1** together with the distribution of terms according to source. The data collection instrument as provided to collaborators is shown in **Table S3**. Each phenotypic category included an ‘Other’ free text field, where submitters could describe additional phenotypes they considered as not listed among the existing terms.

**Table 1:** Distribution of HPO terms in the prospective data collection instrument according to phenotypic category and source.

| Phenotypic Abnormalities (Categories) | <sup>a</sup>FA only | FA & VACTERL-H | FA & Other Sources | FA, VACTERL-H & Other | VACTERL-H only | VACTERL-H & Other | Other Sources only | Total |
| --- | --- | --- | --- | --- | --- | --- | --- | --- |
| Growth | 3 | 1 |  |  |  |  |  | 4 |
| Limbs & Musculoskeletal | 15 |  | 1 | 1 | 5 |  | 6 | 28 |
| Hair, Skin & Nails | 4 |  |  |  |  |  |  | 4 |
| Blood & blood-forming tissues | 4 |  |  |  |  |  | 1 | 5 |
| Head or Neck | 10 |  | 3 | 1 |  |  | 5 | 19 |
| Nervous System | 6 | 3 |  |  | 2 |  | 1 | 12 |
| Eye | 8 | 1 | 2 |  | 3 |  | 1 | 15 |
| Ear | 3 |  |  |  | 3 |  | 1 | 7 |
| Digestive | 5 | 1 |  |  | 3 |  | 5 | 14 |
| Genitourinary | 11 | 1 | 3 | 1 | 2 |  | 9 | 27 |
| Cardiovascular | 9 |  |  |  | 1 |  |  | 10 |
| Respiratory |  | 1 |  |  | 1 |  |  | 2 |
| Prenatal/Birth | 1 |  |  |  | 2 |  |  | 3 |
| Endocrine | 2 |  |  |  |  |  | 1 | 3 |
| Total | 81 | 8 | 9 | 3 | 22 | 0 | 30 | 153 |

HPO – Human Phenotype Ontology. FA=Fanconi anemia, Orphanet code ORPHA:84; VACTERL-H=vertebral anomalies, anal atresia, congenital cardiac disease, tracheoesophageal fistula, renal anomalies, limb defects with hydrocephalus, Orphanet code ORPHA:3412; Other sources are from collaborators and review of publications.
<sup>a</sup> FA only terms listed excludes the following five ORPHA:84 terms: HP:0003220 (Abnormality of chromosome stability), HP:0002863 (Myelodysplastic syndrome), HP:0002664 (Neoplasm/Cancer diagnosis), HP:0000478 (Abnormality of the eye), HP:0001871 (Abnormality of blood and blood-forming tissues). These are very broad phenotypic categories and were collected in more detail or as discrete phenotypic categories on prospective data collection instrument.

### Prospective data collection

Data obtained for 78 FA patients using the data collection instrument was reviewed, following the same variant classification processes as for the retrospective dataset. Five individuals were excluded from analysis due to lack of convincing evidence that one or both variants were (likely) pathogenic, or because no clinical phenotype information was provided. The final prospective dataset included information for 73 FA individuals; 43 were unpublished (one *BRCA1*, 40 *BRCA2* and two *PALB2*), and 30 were previously published with updates provided (all *BRCA2*, information about overlap is provided in **Table S1**). There were 178 phenotypes provided under the ‘Other’ free text field on our data collection instrument. These were reviewed against existing HPO terms in the collection instrument, assigned appropriate HPO terms, and mapped for their relationship to existing and other newly identified terms. The HPO identifier, its relationship to prospective data collection terms, the source, and the number of times reported, are shown for these 178 additional phenotypes in **Table S4**.

### Analysis of phenotypic features

The distribution of the 178 ‘Other’ terms identified in prospective data collection, and their relationship to existing terms was presented graphically, using PhenoScore, an artificial intelligence-based phenomics framework that allows for an efficient rendering of the extensive hierarchical network and can facilitate the identification of potential novel phenotypic connections.^21^ To produce this visualization, Python 3.10 was used to process and display the HPO structure, referencing HPO version 2022-06-11.

The frequency of individual HPO terms observed in our prospectively collected data for FA patients was estimated, and plotted separately for: (i) FA (ORPHA:84) frequency bins designated for these disorders (occasional, 5-29%; frequent, 30-79%; very frequent, 80-99%; (ii) all VACTERL-H (ORPHA:3412) terms; (iii) terms derived from published sources not included in ORPHA:84 or ORPHA:3412. Information was tabulated and shown graphically for descriptive purposes (**Table S5**).

### Comparison of phenotypes reported in published versus prospective datasets

For the 30 FA individuals with both published data and prospectively collected data, information extracted from publications was compared to that collected by phenotyping using the collection instrument, to document the number of additional features captured by prospective data collection for descriptive purposes (**Table S6).**

We then compared the frequencies of FA phenotypes according to phenotypic category for data extracted from publication/s to that collected prospectively for all FA individuals (**Table 2**). For this analysis, the information updates provided for 30 previously published patients were included only in the frequency summaries for the prospective dataset. Information was stratified by patient age at FA diagnosis: ≤5 years (including individuals identified prenatally) compared to >5 years, to align with cancer diagnosis cut-offs set for use of the PM3 code in the ClinGen VCEP specifications for *BRCA1* and *BRCA2*. For reporting and interpretation purposes, the Haldane-Anscombe correction ^22,23^ was applied for comparisons where at least one cell contained zero, and p-values ≤0.05 were considered statistically significant for this descriptive exploratory analysis.

**Table 2:** Frequency of patient characteristics in published vs prospectively collected data according to age at FA diagnosis.

| Phenotypic Abnormalities (Categories) | <sup>a</sup>Age at FA diagnosis ≤5 years |  |  |  |  | <sup>a</sup>Age at FA diagnosis >5 years |  |  |  |  |
| --- | --- | --- | --- | --- | --- | --- | --- | --- | --- | --- |
|  | <sup>b</sup>Published (N=93) |  | <sup>c</sup>Prospective (N=64) |  | <sup>d</sup>p-value | <sup>b</sup>Published (N=35) |  | <sup>c</sup>Prospective (N=9) |  | <sup>d</sup>p-value |
|  | N | % | N | % |  | N | % | N | % |  |
| Growth | 68 | 73.1 | 52 | 81.3 | 0.2 | 13 | 37.1 | 6 | 66.7 | 0.1 |
| Limbs & Musculoskeletal | 53 | 57.0 | 37 | 57.8 | 0.9 | 9 | 25.7 | 4 | 44.4 | 0.3 |
| Hair, Skin & Nails | 58 | 62.4 | 43 | 67.2 | 0.5 | 15 | 42.9 | 7 | 77.8 | 0.06 |
| Blood & blood-forming tissues | 2 | 2.2 | 23 | 35.9 | 1.6x10<sup>-8</sup> | 1 | 2.9 | 5 | 55.6 | 5.0x10<sup>-5</sup> |
| Head or Neck | 36 | 38.7 | 30 | 46.9 | 0.3 | 8 | 22.9 | 1 | 11.1 | 0.4 |
| Nervous System | 50 | 53.8 | 46 | 71.9 | 0.02 | 17 | 48.6 | 4 | 44.4 | 0.8 |
| Eye | 18 | 19.4 | 28 | 43.8 | 0.001 | 4 | 11.4 | 4 | 44.4 | 0.02 |
| Ear | 14 | 15.1 | 26 | 40.6 | 0.0003 | 5 | 14.3 | 4 | 44.4 | 0.05 |
| Digestive | 24 | 25.8 | 24 | 37.5 | 0.1 | 4 | 11.4 | 2 | 22.2 | 0.4 |
| Genitourinary | 32 | 34.4 | 31 | 48.4 | 0.08 | 13 | 37.1 | 4 | 44.4 | 0.7 |
| Cardiovascular | 18 | 19.4 | 18 | 28.1 | 0.2 | 1 | 2.9 | 2 | 22.2 | 0.04 |
| Respiratory | 3 | 3.2 | 4 | 6.3 | 0.4 | 0 | 0.0 | 1 | 11.6 | 0.07 |
| Prenatal/Birth | 1 | 1.1 | 5 | 7.8 | 0.03 | 0 | 0.0 | 0 | 0.0 | 1.0 |
| Endocrine | 4 | 4.3 | 10 | 15.6 | 0.01 | 3 | 8.6 | 2 | 22.2 | 0.3 |
| Patient Cancer diagnosis ≤5 years | 83 | 89.2 | 35 | 54.7 | 9.6x10<sup>-7</sup> | 3 | 8.6 | <sup>e</sup>1 | 11.1 | 0.8 |

d: P-values where contingency tables included N=0 are based on Haldane correction of 0.5 applied to all cells
e: This patient was diagnosed with Wilms tumor at age ≤5 years, but FA diagnosis was not made until after age 5 years.

### Assignment of ACMG/AMP PM3 code for individual variants

For each FA individual considered eligible for phenotypic and genotype analysis (retrospective and prospective patients combined), we reviewed all data collated against the characteristics specified to assign the PM3 code for recessive presentation following ClinGen VCEP specifications for *BRCA1*, *BRCA2* and *PALB2* i.e. chromosome test findings, physical features, pathology findings, cancer diagnosis ≤5yr (Supplemental Information, Supplemental Methods). The rationale for this review was to: (i) determine for how many individuals there was sufficient information to assign PM3 evidence towards pathogenicity for the alleles detected in these individuals with FA; (ii) assess if there might be other features to be considered as important in assigning the PM3 code for FA due to these three genes; (iii) provide a resource for future classification in the context of VCEP activities, including designating which of multiple relatives should be selected as having the most severe presentation for PM3 code application.

### Annotation of predicted and observed variant type and molecular impact for FA-related alleles

For the combined retrospective and prospective datasets (172 individuals), each unique FA-related allele (n=159) was annotated for likely mechanism of impact. As detailed in **Supplemental Data,** annotations included variant location (exon), variant type as a surrogate for primary molecular consequence (synonymous, intronic, missense, deletion, protein termination codon (PTC)), predicted splicing impact, and experimentally-determined variant impact as measured using protein functional and/or splicing assays. Exon numbering matched the exon descriptions of the MANE transcript for *BRCA1* (NM_007294.4), or *BRCA2* (NM_000059.4); i.e. sequential (not legacy) exon numbering was used for *BRCA1*. This included more detailed curation of observed/predicted spliceogenic variants located at the *BRCA2* exon 1 donor motif (**Figure S1**), and the *BRCA2* exon 2 acceptor and donor motifs (**Figure S2**). Aligning with VCEP specifications, variants encoding a PTC were also assessed for susceptibility to nonsense-mediated decay (NMD), and missense substitutions were annotated for location inside or outside a known functional domain. Annotations for unique alleles are shown in **Table S7.** Each variant was then labelled with an annotation acronym capturing combined variant type, splicing prediction, (level) of impact based on experimental data, and NMD prediction for PTC variants (**Table S8).** Exon-level annotation was performed to assess the possibility of rescue of PTC alleles due to single-exon or multi-exon frame (**Table S9**).

In addition, the level and consequence of impact on splicing and/or level of impact on function was tabulated for *BRCA2* FA alleles, specifically to investigate the hypothesis that FA-related alleles may be enriched for hypomorphic variants (**Table S10)**. For this analysis, we defined a hypomorphic allele as a variant with partial impact on mRNA level or protein function, or altered mRNA transcript expression that is expected/shown to result in incomplete impact on function at the protein level, or conflicting results across multiple assays capturing impact on protein function. The initial descriptive analysis was performed using existing functional assay data collated for use as per *BRCA2* VCEP specifications V 1.1, with secondary analysis considering results from another two recently published multiplexed assays of variant effect assessing function for variants in the BRCA2 DNA binding domain.^24,25^

In addition to the experimental impact annotation, graphical representation of allele pairs was undertaken to inform allele severity annotation based on both allele effect and exonic location, separately for each gene. Bi-variant positional schematic plots were generated using R version 4.3.1 with the tidyverse (v2.0.0), ggrepel (v0.9.6) and RColorBrewer (v1.1-3) packages. Final formatting was performed using Inkscape (v0.92.3). Co-ordinates were plotted for a single FA individual per family.

### Annotation of allele and genotype severity score for assessing association with clinical presentation

Previous findings from Radulovic et al showed that cancer age at diagnosis in *BRCA2* FA individuals was associated with allele severity impact, defined based on: variant type, location ≤ exon 11 or > exon 11, or splicing prediction score changes for splice site dinucleotide variants.^26^ Building on this approach, we recreated an allele scoring system similar to that of Radulovic et al,^26^ and also created a novel *BRCA2* allele severity scoring system, that included several permutations of severity score based on variant type, predicted/observed molecular impact, location, protein stability, and possible rescue due to in-frame exon skipping or possible upregulation of known alternative in-frame isoforms (**Table S7, Table S9**). See **Supplemental Methods** for details. Using labels as defined by Radulovic et al, scores for individual alleles ranged from 0 (most severe impact) to 2 (least severe impact).

We then conducted Kaplan-Meier survival analysis and Cox regression modelling to investigate the impact of genotype severity score (i.e. allele 1 severity score + allele 2 severity score) on age at cancer diagnosis in *BRCA2* FA individuals. For this analysis, prenatal FA individuals were excluded, as was a single cancer-affected case with unknown age at diagnosis. Individuals affected with cancer were censored as affected at age of diagnosis, and individuals with benign tumors or cancer-unaffected were censored at age of death, or last known review. Survival analysis and Cox regression modelling were performed using STATA SE v.15 (Stata Corp., USA). **Table S1** provides more details on assumptions made regarding inferred age at presentation/review for a subset of cases.

The best fitting Cox regression model for association of *BRCA2* genotype score with age at cancer diagnosis was selected for additional analysis that separated genotype score 2 according to the component allele severity scores (0+2 vs 1+1). The allele severity scoring system that resulted in the best fitting *BRCA2* allele severity model was then applied to derive allele and genotype severity scores for *BRCA1* and *PALB2* FA individuals for descriptive analysis.

### Case-control analysis estimating risk of breast cancer by allele severity score

Building on findings showing association of combined allele severity score with age at cancer onset, we assessed the risk of breast cancer associated with heterozygous *BRCA1* and *BRCA2* pathogenic variant status, stratified by allele severity score assigned using the same approach as for FA-related alleles (see **Supplemental Methods** for more details). Following the approach applied for Cox regression analysis of FA *BRCA2* individuals, analysis was performed considering different permutations of allele severity score. Burden analyses in which odds ratios and 95% confidence intervals for breast cancer associated with the presence of any variant in a given allele severity score category were estimated by means of logistic regression in a combined dataset of 96,991 female breast cancer cases and 302,116 unaffected controls aggregated from the BRIDGES study of the Breast Cancer Association Consortium (BCAC),^27^ the Cancer Risk Estimates Related to Susceptibility (CARRIERS) consortium^28^ and the UK Biobank (UKB).^29,30^ Dataset quality control and filtering criteria were described elsewhere.^31^ Associations were adjusted for age and study country for the BCAC dataset, age and ethnic group for the CARRIERS dataset, and age and genetic ancestry for the UKB dataset. Odds ratios and standard errors estimated from each dataset were combined in a fixed-effects, inverse-variance meta-analysis using the ‘metafor’ R package to derive an overall test of association.

## RESULTS AND DISCUSSION

### Phenotypes observed in prospective data collection

Overall, 296 features were observed in FA individuals included in the prospective cohort. The number of observations of each HPO term is shown in **Table S4**. The features observed included 82 of 104 ORPHA:84 terms, 14 of 22 terms originating only from the ORPHA:3412 VACTERL list, and 20 of 30 terms sourced from publications or collaborators. Of the 178 phenotypes provided as free text, simplistic relationship mapping established that 84 terms could be related by hierarchy to other features listed in the data collection instrument: 68 terms were descendants of existing terms (24 child, 29 grandchild, 11 great grandchild, and 4 great-great grandchild); two terms were grandparents of existing terms; and 15 terms were in the same HPO hierarchy as existing terms (designated ‘sibling’ terms). The remaining 94 terms were considered to be new terms not directly linked to HPO terms listed in the data collection instrument (called “new” terms for display purposes).

The distribution of ‘Other’ terms across phenotypic categories, and their relationship mapped to existing terms in the prospective data collection sheet, is shown in **Figure 3**. The most frequently reported ‘Other’ phenotypes were: short thumb (also reported as thumb hypoplasia/brachydactyly, n=10); failure to thrive/small for gestational age (n=6); pelvic kidney (n=6); posteriorly rotated ears (n=4); stenosis of the external auditory canal (n=4); and vesicoureteric reflux (n=4). Ten terms (sacral dimple, preaxial polydactyly, bone marrow hypocellularity, convex nasal ridge, holoprosencephaly, bilateral sensorineural hearing impairment, fused or horseshoe kidney, ventricular septal defect, growth hormone deficiency, small/ectopic pituitary) were each reported three times, and another 26 terms were each reported twice.

**Figure 3:**
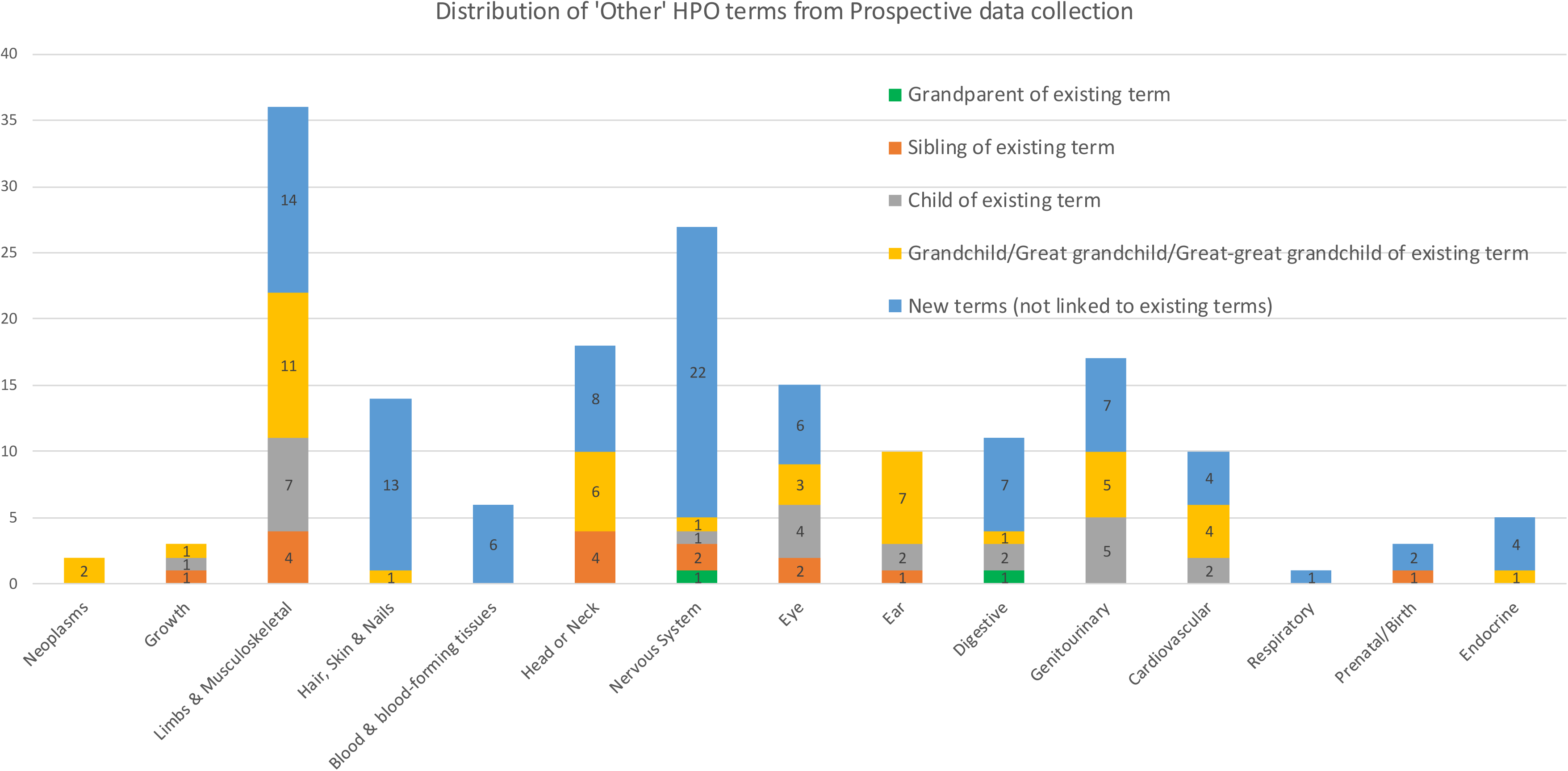
Distribution of ‘Other’ Human Phenotype Ontology terms obtained in prospective data collection. The 178 ‘Other’ terms were distributed across phenotypic categories as follows: Neoplasms (n=2, specified by histology); Growth (n=3); Limbs and Musculoskeletal (n=36); Hair, Skin and Nails (n=14); Blood and Blood-forming tissues (n=6); Head or Neck (n=19); Nervous System (n=27); Eye (n=15); Ear (n=10); Digestive (n=11); Genitourinary (n=17); Cardiovascular (n=10); Respiratory (n=1); Prenatal/Birth (n=3); Endocrine (n=5).

These findings are apparent from the network visualization (**Figure 4, Figure S3**), showing that the new terms can be ontologically connected to existing FA HPO terms, albeit with many new terms presenting as outliers in the hierarchical connection (**Figure S3**). These findings indicate the need to redefine the existing list of FA-related HPO terms using an ontogenic phenomics approach, and scope to extend this list to include several new features in the future.

**Figure 4.**
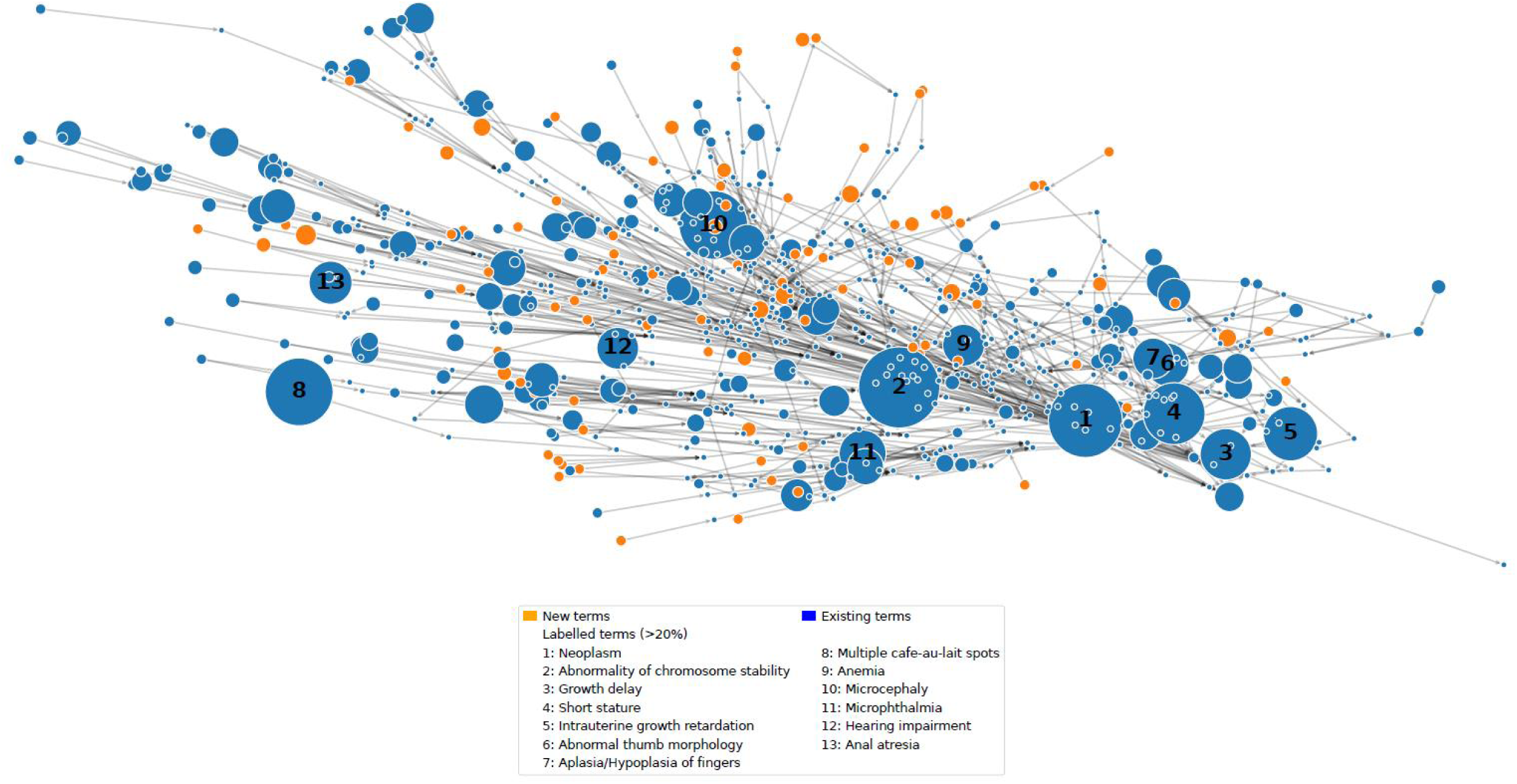
Overview of Phenotypic Data in Human Phenotype Ontology Terms. This network visualization illustrates the distribution and relationships of phenotypic terms as defined by the Human Phenotype Ontology (HPO). Each circle (node) represents an HPO term, and edges (grey lines) indicate hierarchical or ontological connections among those terms, following the hierarchy of the HPO. Two key categories of terms are displayed: (1) New terms (orange nodes): these are phenotypic features previously not reported as related to Fanconi anemia, and not directly linked to HPO terms in the data collection instrument; (2) Existing terms (blue nodes): well-established HPO terms and those related by hierarchy. Node size reflects the relative frequency of a particular phenotype in the dataset—larger circles denote phenotypes observed more frequently among the subjects studied. The legend lists the specific phenotypic features representing >20% of observations, with numbers for each of these features shown on the network

**Figure 5:**
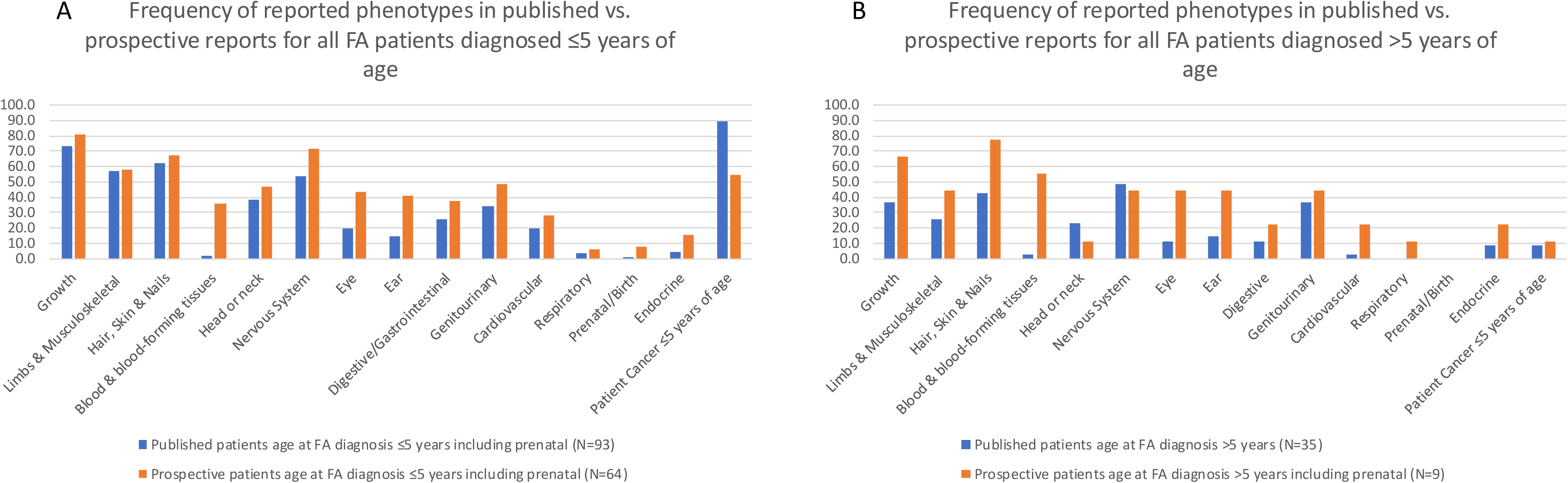
Comparison between frequency of reported phenotypes in published versus prospective reports for FA individuals according to phenotypic category. Y axis shows % of individuals reported to have one characteristic in a given phenotypic category. Figure 5A compares the frequency of phenotypes among individuals diagnosed with FA under age 5 years (including prenatal FA individuals), between published (n=93), and prospective (n=64) FA individuals. Figure 5B compares the frequency of phenotypes among individuals diagnosed with FA over age 5 between published (n=35) and prospective (n=9) FA individuals. See **Table 2** for details of statistical comparisons between groups.

### Frequency of ORPHA:84 and ORPHA:3412 HPO terms observed in the prospective dataset of *BRCA1*, *BRCA2,* and *PALB2* FA individuals

The HPO terms classified as ‘very frequent’ (80-99%) for FA (ORPHA:84) ranged in frequency in our prospective data from 0-81.2%; ‘frequent’ (30-79%) ranged from 1.4- 68.5%; and ‘occasional’ (5-29%) ranged from 0-56.2% after correction for sex where relevant (**Table S5a – 5c**). HPO terms unique to VACTERL-H (ORPHA:3412) that were classified as ‘very frequent’ ranged in our dataset from 0-5.3%, whereas ‘frequent’ ranged from 0-10.7%, and ‘occasional’ ranged from 0-12% (**Table S5d**). HPO terms added from FA-related publications^18–20^ and personal communications with collaborators ranged in frequency from 0-23.5% (**Table S5e**).

### Comparison of phenotypic data reported in publications versus prospective collection

For 27 of the 30 individuals with FA and for which data was sourced from existing sources, and via prospective collection, more features were provided through the standardized prospective collection compared to extraction from publications (**Table S6**). For several individuals, the additional phenotype data led to altered assignment of the PM3 code, for use in variant curation.

Compared to data extracted from published studies (**Table 2)**, prospective reports for FA patients diagnosed ≤5 years of age had significantly higher frequencies of abnormalities in Blood and blood-forming tissues (36% vs 2%; p=1.6×10^-8^), Nervous system (72% vs 54%; p=0.02), Eye (44% vs 19%, p=0.001), Ear (41% vs 15%; p=0.0003), Prenatal/Birth (8% vs 1%; p=0.03) and Endocrine (16% vs 4%; p=0.01). Conversely, published studies were significantly more likely than prospective studies to report individuals diagnosed with FA ≤5 years of age who also had at least one cancer diagnosis (89% vs 55%; p=9.6×10^-7^). In the subset of individuals diagnosed with FA >5 years of age, prospective reports had significantly higher frequencies of abnormalities of Blood and blood forming tissues than published reports (56% vs 3%; p=5.0×10^-5^), Eye (44% vs 11%; p=0.02), Ear (44% vs 14%; p=0.05) and Cardiovascular abnormalities (22% vs 3%; p=0.04). These differences likely reflect biases in case studies identified as well as biases in the associated information selected for presentation in published reports of FA individuals e.g. there was no or poor annotation of non-cancer hematological features in most publications.

### Alignment of recessive presentation features with VCEP specifications

Review of each individual with FA for the VCEP-specified characteristics designated to assign the PM3 recessive code is captured in **Table S1**. The 172 individuals with FA represented 140 different families. Of the nine individuals with prenatal presentation, three met the PM3 criterion, while characteristics reported for the remainder was insufficient to assign PM3 - all presented with physical features of FA, but were (apparently) untested for chromosomal breakage, and had no information reported relevant to pathology findings.

For the remaining 163 individuals, PM3 was met by 146 individuals representing 121 families; considering only code assignment per family, the code was met by 107 individuals at moderate weight, and 14 at supporting weight. For the remaining individuals, PM3 code was not met largely because of insufficient information. After considering censoring age due to death, only three individuals had presentation considered “not characteristic” as defined by the VCEP specifications: one individual with positive chromosome test results, breast cancer diagnosis in age range 21-25 years, and no reported physical or pathology features;^32^ one individual with negative chromosome test results, and colorectal cancer diagnosis in age range 36-40 years;^33^ and one individual with negative chromosome test results, mild physical features, and toxicity associated with treatment for breast cancer diagnosis in age range 26-30 years.^34^ Notably, the latter individual was biallelic for the established high-risk pathogenic allele *BRCA1* c.181T>G p.(Cys61Gly) and the known pathogenic allele *BRCA1* c.5096G>A p.(Arg1699Gln)^34^ confirmed to be associated with reduced penetrance^35,36^.

### Overview of predicted and observed variant type and molecular impact for FA-related alleles

The combined dataset of non-overlapping *BRCA1*, *BRCA2* or *PALB2* FA individuals identified from published or unpublished data, after exclusions, included 11 individuals with FA due to *BRCA1* (from 9 distinct families), 144 individuals with FA due to *BRCA2* (from 115 families) and 17 individuals with FA due to *PALB2* (from 15 families) (**Table 3**). Many of the FA-related alleles were observed multiple times across different families. Specifically, 15 unique *BRCA1* alleles observed in 9 *BRCA1* families, 122 unique *BRCA2* alleles in 115 *BRCA2* families, and 22 unique *PALB2* alleles in the 15 *PALB2* families.

**Table 3:** Distribution of FA-related alleles according to predicted and observed variant impact on function.

|  | *BRCA1* | *BRCA2* | *PALB2* |
| --- | --- | --- | --- |
| <sup>a</sup> Total number of FA individuals | 11 | 144 | 17 |
| *Published only* | 10 | 74 | 15 |
| *Published updated* | 0 | 30 | 0 |
| *Previously unpublished* | 1 | 40 | 2 |
| Total number of families | 9 | 115 | 15 |

| <sup>b</sup> Variant Annotation Description | Unique *BRCA1* alleles | <sup>c</sup> Allele observations | Unique *BRCA2* alleles | <sup>c</sup> Allele observations | Unique *PALB2* alleles | <sup>c</sup> Allele observations |
| --- | --- | --- | --- | --- | --- | --- |
| *(Total n)* | 15 | 18 | 122 | 230 | 22 | 30 |
| Synonymous/Intronic splicing predicted/proven | 0 | 0 | 6 | 13 | 3 | 4 |
| Missense/in-frame deletion within functional domain | 6 | 6 | 19 | 29 | 0 | 0 |
| Missense in domain splicing predicted & proven | 0 | 0 | 2 | 3 | 0 | 0 |
| Missense out domain splicing predicted & proven | 0 | 0 | 4 | 15 | 0 | 0 |
| <sup>d</sup> Large deletion | 0 | 0 | 1 | 1 | 1 | 1 |
| <sup>d</sup> PTC | 8 | 11 | 69 | 109 | 10 | 13 |
| <sup>d</sup> PTC no NMD | 0 | 0 | 3 | 14 | 4 | 8 |
| <sup>d</sup> PTC splicing predicted/proven | 1 | 1 | 3 | 23 | 3 | 3 |
| <sup>d</sup> Intronic Splice Site dinucleotide predicted loss-of-function as per PVS1 decision tree | 0 | 0 | 11 | 19 | 1 | 1 |
| Intronic Splice Site dinucleotide predicted/proven indel or regulatory | 0 | 0 | 4 | 4 | 0 | 0 |
| <sup>d</sup> Proportion of variants with (predicted) impact considered loss-of-function as per ClinGen PVS1 decision tree | 0.60 | 0.67 | 0.71 | 0.72 | 0.86 | 0.87 |

<sup>a</sup> Includes only individuals with minimum data to be included in the phenotype-genotype component of the study (P, PU and U codes, Table S1).
<sup>b</sup> See Supplemental Information and Table S7 for more details on variant annotation and categorization. A *BRCA1* small in-frame deletion was coded as missense for presentation.
<sup>c</sup> Total observations across unrelated families (i.e. excluding relatives).
<sup>d</sup> Variant types considered assessable via a PVS1 decision tree, as detailed in Abou Tayoun et al<sup>37</sup>.

Alleles generally considered to be loss-of-function based on variant type (PTC, splice site dinucleotide, and deletion, as assessed using a PVS1 decision tree^37^) comprised the majority of unique alleles across the three genes (*BRCA1*, 60%; *BRCA2* 71%, *PALB2* 86%), and also the majority of all alleles observed per gene in the unrelated families (≥ 67%). This included three unique *BRCA2* and four unique *PALB2* PTC variants predicted to escape NMD but which would still result in loss of critical functional residues. The five *BRCA1* unique variants encoding a missense substitution, plus one in-frame deletion variant, all fell within a known clinically important functional domain (two in the RING domain, the remainder in the BRCA1 C-terminal domain). For *BRCA2*, 21 predicted missense substitution variants fell within the DNA binding domain, two of which had mechanism of impact through splicing; the remaining four “missense” variants located outside of a functional domain all impacted splicing.

A summary of experimental data indicating level of impact on splicing and/or (protein) function for *BRCA2* FA alleles is shown in **Table S10**, with a more detailed explanation of the findings reported in **Supplemental Results**. In brief, experimental data was available for all *BRCA2* missense and synonymous variants, all intronic variants located outside of the splice site dinucleotide positions, and half of the intronic variants located at the dinucleotide positions. Excluding variants encoding a PTC, 16% (6/38) of those tested experimentally to capture the allele molecular effect (protein and/or splicing) showed partial or conflicting functional or splicing impact based on one or more assays. In comparison, review of calibrated functional assay results compiled for use in VCEP curation under the updated Specifications V 1.2 (https://cspec.genome.network/cspec/ui/svi/doc/GN097) showed that impact was partial or conflicting for 29% (80/270) of the BRCA2 missense variants recorded as having any impact (partial or complete) from at least one study. This simplistic comparison suggested that the proportion of variants with partial or conflicting impact on function versus complete impact is not obviously greater for alleles observed in *BRCA2* FA individuals compared to the pool of variants with assay results. Additionally, when considering findings from two recently published multiplexed assays of variant effect (MAVE) capturing the functional impact at the mRNA and protein levels,^24,25^ yet to be formally included in VCEP specifications, further inconsistencies were introduced. These discrepancies were due to a difference in functional impact for one or both of the MAVE studies compared to those drawn from existing functional data (24 variants), or differences in results between the two MAVE studies. Overall this observation highlights a limitation in using “lack of agreement between functional assays” to infer whether a variant may potentially have partial impact on function, since different functional assays can measure different aspects of function, and also because inconsistency in functional impact results for individual variants may reasonably be expected to increase as more assay findings (and thus more experimental error) are included in the annotations.

### Within-gene distribution of biallelic pairs observed in individuals with FA

Graphical representation of allele pairs is shown in **Figure S4** *(BRCA1*), **Figure S5** (*BRCA2*), and **Figure S6** (*PALB2*). The majority of *BRCA1* pairs have one or both alleles located in exon 10, consistent with the hypothesis that upregulation of naturally occurring in-frame splicing may be a key factor acting to rescue lethality for *BRCA1* biallelic pathogenic variant status.^12,38^ The remaining three pairs included at least one missense variant, including the *BRCA1* c.5096G>A p.(Arg1699Gln) variant demonstrating ambiguous findings across different functional assays, and confirmed to be associated with reduced breast and ovarian cancer risk compared to the average *BRCA1* PTC pathogenic variant.^35,36^ For *BRCA2*, the most notable observation is the paucity of biallelic variants in the largest exon (exon 11).

Only one individual was bi-allelic for pathogenic variants in this exon, homozygote for *BRCA2* c.3751dup p.(Thr1251Asnfs*14), diagnosed after pregnancy termination at 13 weeks. For *PALB2*, it was notable that six of the 11 individuals with FA had at least one PTC allele predicted to escape NMD.

### Association of *BRCA2* genotype severity score with age at cancer diagnosis

Cox regression analysis was undertaken to assess the association between age at cancer diagnosis and genotype severity score (based on the summed allele severity score), expanding previously reported findings by Radulovic et al.^26^ Analysis using an allele scoring system aligned with that of Radulovic et al ^26^ showed a significant association with age at cancer onset (**Figure S7**, Chisq (4df)=18.04, p=0.001). The result was slightly more significant using the baseline severity score system selected for this study (**Figure S8**, Chisq (4df)=19.33, p=0.0007). Association results from baseline and several permutations of this severity scoring system are shown in **Table S11**. The best fitting model (**Figure 6**) was as follows: severity score of missense variants was based on predicted protein instability rather than functional assay data, a less severe score was applied to splice variants with partial impact on splicing or resulting in large in-frame transcripts that do not delete a known functional domain, and a less severe score was applied to PTC variants located in exons with potential for rescue due to single or multi-exon in-frame skipping. When genotype severity score was categorized into five groups from most to least severe (0, 1, 2, 3, 4), age at cancer onset was significantly later for individuals with genotype score 1 (Hazard Ratio (HR) 0.46), score 2 (HR 0.23), score 3 (HR 0.13) and score 4 (HR 0.11) compared to those with genotype severity score 0 (Chisq (4df) = 40.94, Log rank test p=2.8 x 10^-8^). The strength of association was similar (Chisq (3df) = 41.81, Log rank test p=1.8 x 10^-8^) when considering the component allele severity scores that contributed to the genotype scores (**Figure 7**). The HR was unchanged for allele score 0+1 (i.e. genotype severity score 1), and was marginally different (0.25) for the subset of individuals with genotype severity score 2 due to component allele scores 1+1. The HR was 0.13 for the pool of remaining genotype combinations, each comprised of at least one allele score 2 (allele score 2+0, 2+1, and 2+2).

**Figure 6:**
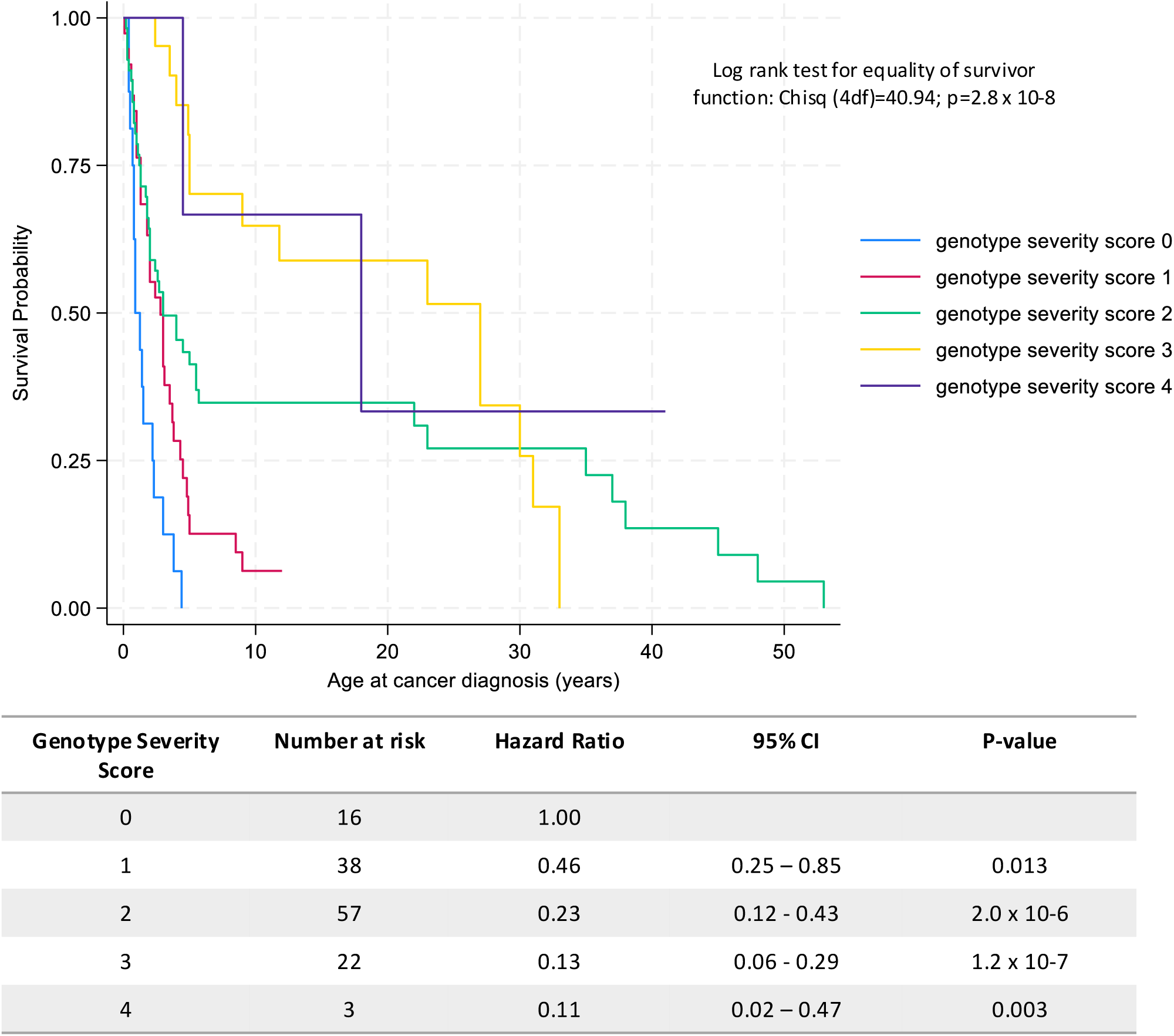
Kaplan-Meier probabilities and Cox regression analysis assessing association of genotype severity score with age at diagnosis of cancer for *BRCA2* FA individuals, for best-fitting genotype severity score model. For this model, the allele severity scores were based on: PTC (noNMD) score 2; PTC exon 10, 11, 12 score 1 (exon rescue); PTC exon 4-7 score 1 (potential multi-exon rescue); PTC exon 14 score 1 (potential escape from NMD due to large exon size); other PTC score 0; Splice site dinucleotide with regulatory impact score 1; splicing complete impact and observed transcript expected to under NMD score 0; splicing complete impact and transcript encodes larger in-frame deleted expected to escape NMD score 1; splicing partial impact score 1; missense outside domain (all with splice impact) score 0; missense in domain predicted unstable score 1; missense in domain predicted stable score 2. See Table S7 for further description of the annotations informing allele severity scores. Survival plots and Cox proportional hazard Ratios are estimated, using genotype severity score 0 as reference, for severity scores 1, 2, 3 and 4.

**Figure 7.**
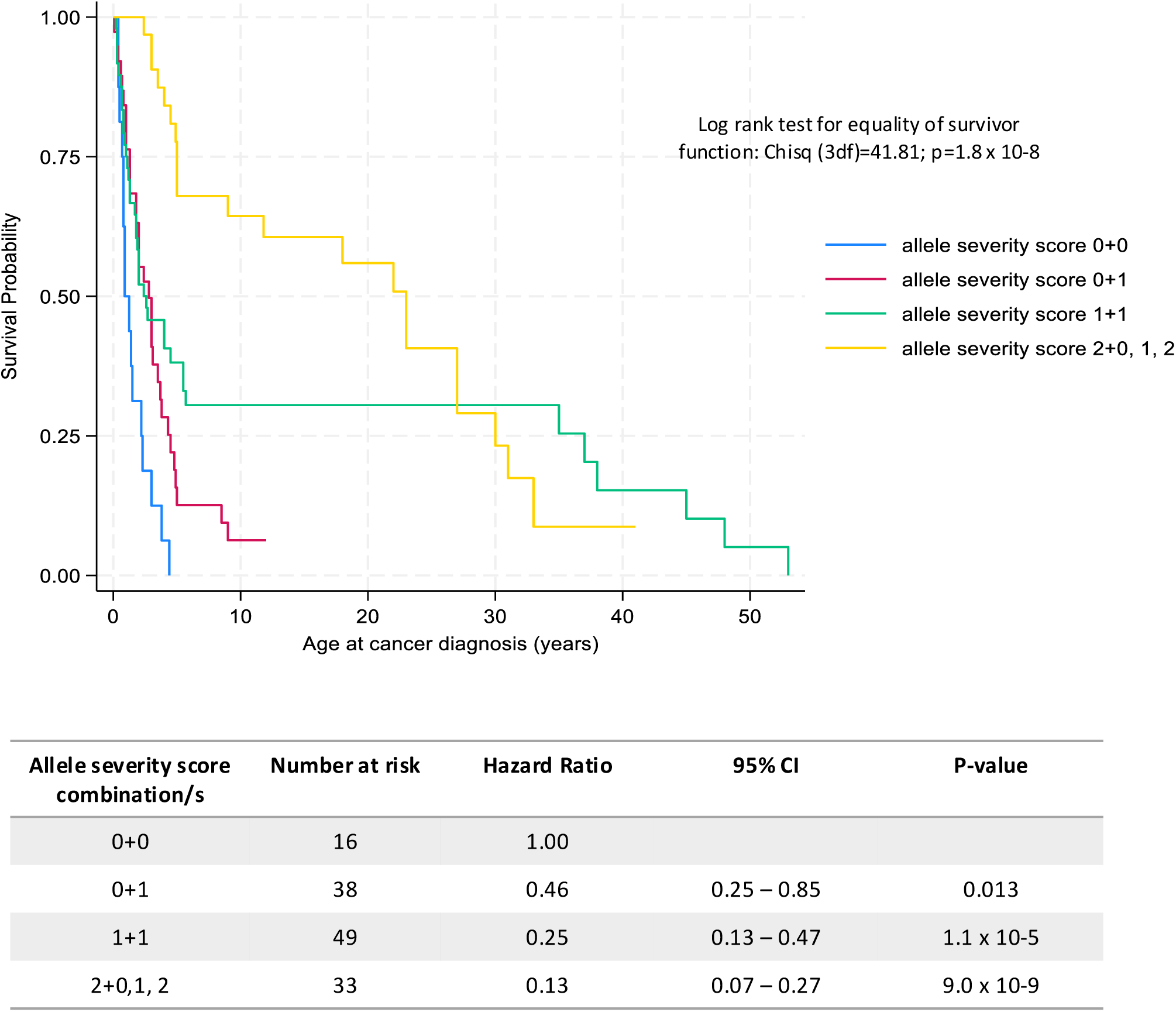
Kaplan-Meier probabilities and Cox regression analysis assessing association of genotype severity score with age at diagnosis of cancer for *BRCA2* FA individuals, considering allele score components of genotype severity. For this model, the allele severity scores were applied as for best-fitting genotype severity model shown in Figure 6, but genotype severity was grouped to consider the component allele severity score, as follows: genotype score 0, all based on allele severity 0+0; genotype score 1, all based on allele severity 0+1; genotype score 2 based on allele severity 1+1; genotype score 2 or greater (score 2, 3, or 4) where at least one allele was coded with severity score 2, and the remaining allele was score 0 or 1 or 2. Survival plots and Cox proportional hazard Ratios are estimated, using genotype severity score 0 as reference.

### Association of *BRCA1* and *PALB2* genotype severity score with FA phenotype

The allele severity scoring system that yielded the best fitting *BRCA2* allele severity model was then applied to derive allele and genotype severity scores for individuals with FA due to *BRCA1* and *PALB2*. Comparisons of the relative proportions of alleles by severity score for each gene, are shown in **Table S12**. Although these comparisons were based on small numbers, it was notable that no *BRCA1* alleles were assigned a severity score of 0 (i.e., PTC variants or splicing alleles with no rescue expected), and that most (72%) alleles were assigned score of 2, with most of these being PTC variants with potential rescue by in-frame isoform expression, and potential escape from NMD due to large exon size (**Table S7**). This observation is consistent with the hypothesis that individuals bi-allelic for BRCA1 pathogenic variants can *only* survive if both variant retain some function. In contrast, FA-related alleles in *PALB2* were distributed across all three severity score categories, as already observed for *BRCA2*.

Clinical presentation age and features, grouped into broad categories for simplicity, showed no obvious pattern in relation to *BRCA1* or *PALB2* genotype severity scores, or the component allele scores (**Table S13**). Genotype score distribution was consistent with the distribution of individual allele severity scores for the different genes. While most *BRCA1* FA individuals (nine out of 11) had genotype severity score of 3 or more, genotype severity score 2 was assigned for the single prenatal case age, and for two individuals with relatively late age at cancer diagnosis (in age ranges 21-25 years, and 26-30 years). Almost half (47%) of *PALB2*-related FA individuals had a genotype severity score of 1, including the single prenatal case. Genotype severity score 2 was assigned to 42% of *BRCA2*-related FA individuals, including four of seven individuals with prenatal presentation. Additionally, comparison of the phenotype for pairs or trios of siblings across 30 families (**Table S14**) revealed differences in age at cancer diagnosis (categorized as ≤ vs > 5 years) for both of two *BRCA1* families (genotype severity score 4), one of two *PALB2* families (with genotype severity score 1), and for three of 28 *BRCA2* families (with genotype (allele) severity scores of 1 (0+1), 2 (0+2) and 3 (1+2)).

### Association of *BRCA1* and *BRCA2* allele severity score with risk of breast cancer

Results from meta-analysis of three breast cancer case-control datasets is shown in **Table 4**, for different allele severity score options, with results for each dataset detailed in **Table S15** (*BRCA1*) and **Table S16** (*BRCA2*). For *BRCA1*, the baseline severity scoring option showed the best separation of effect (Pcontinuous=7.11×10^-153^, **Table S15**), with alleles annotated as severity score 2 demonstrating a reduced breast cancer risk (OR=4.88, 95% confidence interval (CI) 2.89-8.26; P=3.34×10^-9^), compared to those with severity score 0 (OR=11.36, 95% CI 8.51-15.17) and severity score 1 (OR=11.06, 95% CI 8.86-13.80). Applying allele scoring options A, B and C led to few changes in allele severity coding for unique alleles. In contrast, further upweighting PTC variants within in-frame exon 10 (Option D) due to evidence for possible rescue by naturally occurring splicing changed allele score annotation for a large proportion of variants/individuals, and there was large overlap in confidence intervals around the risk estimates for the three allele severity score categories. That is, of the allele scoring options assessed in this study, the baseline scoring approach provided the best model for predicting differences in risk for *BRCA1* allele effect and location. Although the CIs around the OR estimates for this baseline model indicated a distinctly lower risk associated with *BRCA1* alleles of severity score 2 compared to those with score 1 or 0, the risk estimate itself falls into the range (OR>4.0) considered “high risk” for breast cancer^39^, and is thus unlikely to inform changes in clinical management for *BRCA1* pathogenic variant heterozygotes in relation to breast cancer risk.

**Table 4:** Risk of breast cancer according to allele severity score - summary results from meta-analysis of three datasets^a^.

| Gene | Allele Severity Option | Allele Severity Scoring description | Allele Severity Score | Total unique variants (n) | Case Het (n) | Control Het (n) | OR (95% CI) | P |
| --- | --- | --- | --- | --- | --- | --- | --- | --- |
| *BRCA1* | *Baseline* | PTC (noNMD) score 2; PTC exon 10 score 1 (in-frame exon rescue); other PTC score 0. Impacts start site score 0. Splicing complete impact score 0, partial impact score 1. Missense in domain complete functional impact score 1, partial or conflicting impact score 2. | 0 | 114 | 362 | 64 | 11.36 (8.51–15.17) | 5.92E-61 |
|  |  |  | 1 | 205 | 531 | 124 | 11.06 (8.86–13.80) | 1.44E-100 |
|  |  |  | 2 | 13 | 48 | 26 | 4.88 (2.89–8.26) | 3.34E-09 |
|  | *A* | As for baseline, but: (i) replace missense functional impact score with missense stability score (unstable score 1, stable score 2). | 0 | 114 | 362 | 64 | 11.36 (8.51–15.17) | 5.92E-61 |
|  |  |  | 1 | 205 | 528 | 123 | 10.94 (8.75–13.67) | 2.20E-98 |
|  |  |  | 2 | 13 | 51 | 27 | 5.46 (3.30–9.02) | 3.87E-11 |
|  | *B* | As for baseline, but: (i) replace missense functional impact score with missense stability score (unstable score 1, stable score 2); (ii) apply score 1 for subset of splicing variants that lead to transcripts encoding a larger in-frame deletion expected to escape NMD. | 0 | 108 | 351 | 61 | 11.34 (8.44–15.23) | 1.75E-58 |
|  |  |  | 1 | 209 | 527 | 125 | 10.87 (8.71–13.56) | 5.26E-99 |
|  |  |  | 2 | 15 | 63 | 28 | 6.15 (3.78–9.99) | 2.338E-13 |
|  | *C* | As for baseline, but: (i) replace missense functional impact score with missense stability score (unstable score 1, stable score 2); (ii) upweight score for subset of splicing variants that lead to transcripts encoding a larger in-frame deletion expected to escape NMD; (iii) apply score 1 for PTC variants within *BRCA1* exon 8 and 9 due to potential multi-exon in-frame rescue | 0 | 106 | 350 | 60 | 11.47 (8.53–15.42) | 1.23E-58 |
|  |  |  | 1 | 211 | 528 | 126 | 10.80 (8.66–13.47) | 9.10E-99 |
|  |  |  | 2 | 15 | 63 | 28 | 6.15 (3.78–9.99) | 2.33E-13 |
|  | *D* | As for baseline, but: (i) replace missense functional impact score with missense stability score (unstable score 1, stable score 2); (ii) upweight score for subset of splicing variants that lead to transcripts encoding a larger in-frame deletion expected to escape NMD; (iii) apply score 1 for PTC variants within *BRCA1* exon 8 and 9 due to potential multi-exon in-frame rescue; (iv), upweight PTC variants in *BRCA1* exon 10 due to additional molecular evidence for in-frame rescue. | 0 | 106 | 350 | 60 | 11.47 (8.53–15.42) | 1.24E-58 |
|  |  |  | 1 | 34 | 106 | 17 | 13.98 (7.39–26.46) | 5.23E-16 |
|  |  |  | 2 | 192 | 485 | 137 | 9.40 (7.60–11.61) | 2.45E-95 |
| *BRCA2* | *Baseline* | PTC (noNMD) score 2; PTC exon 10, 11, 12 score 1 (exon rescue); other PTC score 0. Splice site with regulatory impact score 1. Splicing complete impact score 0, partial impact score 1. Missense outside domain (all with splice impact) score 0. Missense in domain complete functional impact score 1, partial or conflicting impact score 2. | 0 | 235 | 452 | 158 | 8.45 (6.85–10.43) | 8.96E-88 |
|  |  |  | 1 | 391 | 1,014 | 585 | 5.33 (4.72–6.01) | 2.07E-163 |
|  |  |  | 2 | 7 | 16 | 46 | 1.26 (0.58–2.72) | 5.56E-01 |
|  | *A* | As for baseline, but: (i) replace missense functional impact score with missense stability score (unstable score 1, stable score 2). | 0 | 235 | 452 | 158 | 8.45 (6.85–10.43) | 8.96E-88 |
|  |  |  | 1 | 382 | 970 | 561 | 5.35 (4.73–6.05) | 2.64E-156 |
|  |  |  | 2 | 16 | 60 | 70 | 2.64 (1.73–4.04) | 6.97E-06 |
|  | *B* | As for baseline, but: (i) replace missense functional impact score with missense stability score (unstable score 1, stable score 2); (ii) apply score 1 for subset of splicing variants that lead to transcripts encoding a larger in-frame deletion expected to escape NMD. | 0 | 226 | 444 | 152 | 8.78 (7.08–10.88) | 9.68E-88 |
|  |  |  | 1 | 390 | 978 | 567 | 5.30 (4.69–5.99) | 3.75E-156 |
|  |  |  | 2 | 17 | 61 | 70 | 2.66 (1.74–4.07) | 5.71E-06 |
|  | C | As for baseline, but: (i) replace missense functional impact score with missense stability score (unstable score 1, stable score 2); (ii) apply score 1 for subset of splicing variants that lead to transcripts encoding a larger in-frame deletion expected to escape NMD; (iii) apply score 1 for PTC and missense variants within *BRCA2* exon 4-7 due to potential multi-exon in-frame rescue. | 0 | 203 | 404 | 127 | 9.59 (7.64–12.05) | 5.43E-84 |
|  |  |  | 1 | 413 | 1,018 | 592 | 5.27 (4.67–5.94) | 2.97E-160 |
|  |  |  | 2 | 17 | 61 | 70 | 2.66 (1.74–4.07) | 5.69E-06 |
|  | D | As for baseline, but: (i) replace missense functional impact score with missense stability score (unstable score 1, stable score 2); (ii) apply score 1 for subset of splicing variants that lead to transcripts encoding a larger in-frame deletion expected to escape NMD; (iii) apply score 1 for PTC and missense variants within *BRCA2* exon 4-7 due to potential multi-exon rescue; (iv) apply score 1 for PTC in exon 14 due to exon size and potential to escape NMD. Allele scoring method as for best-fitting *BRCA2* FA genotype severity score model. | 0 | 186 | 384 | 121 | 9.37 (7.41–11.85) | 6.65E-78 |
|  |  |  | 1 | 430 | 1,037 | 599 | 5.34 (4.74–6.02) | 6.72E-166 |
|  |  |  | 2 | 17 | 61 | 70 | 2.66 (1.74–4.07) | 5.69E-06 |

<sup>a</sup> Results for individual datasets are provided in Table S15 (*BRCA1*) and Table S16 (*BRCA2*). Total n: cases = 96691, controls = 302116. PTC, protein termination codon; NMD, nonsense-mediated decay; Het, Heterozygote; OR, Odds Ratio; CI, Confidence Interval.

For *BRCA2*, all allele severity score options revealed differences in magnitude of risk for alleles with severity score 0 (alleles annotated as most severe impact, OR range 8.45 to 9.57) compared to those with severity score 1 (OR range 5.30 to 5.34), with non-overlapping CIs for within-option comparisons. For alleles with severity score 2 (least severe impact), there the risk estimate was below 3, ranging from a non-significant 1.3-fold risk (0.58-2.71) for baseline annotation, to OR 2.64 (1.72-4.03) for Option A, or OR 2.66 (1.74-4.07) for all the remaining score options. Based on the continuous test (**Table S16**), there was no obviously better separation of effect for one scoring option method over another. For allele severity scoring option D, equivalent to the severity score model with best fit for association with age at cancer onset in *BRCA2* FA individuals, the magnitude of breast cancer risk differed according allele score, as follows: score 0, OR 9.37 (95% CI 7.41-11.85), score 1, OR 5.34 (95% CI 4.74-6.02), and score 2, OR 2.66 (95% CI 1.74-4.07). Overall, these findings indicate that this allele severity scoring system, or perhaps an improved derivation thereof, could potentially be used as an indicator to highlight individual variants as suspected reduced penetrance *BRCA2* alleles, with implications for breast cancer risk management stratification of individuals heterozygous for a *BRCA2* pathogenic variant.

## GENERAL DISCUSSION AND CONCLUSIONS

Using a comprehensive data collection instrument to record features observed in patients with FA due to *BRCA1*, *BRCA2* or *PALB2* pathogenic variants, our study results support the need to expand the list of FA-related features to include additional phenotypes. This includes need for flexibility to incorporate additional HPO terms that span the grandparent-parent-child- grandchild hierarchy of existing FA-related terms (ORPHA:84), and possible expansion to record features captured by multiple HPO terms unique to VACTERL-H (ORPHA:3412), or drawn from the literature, at frequency ranging from 5 to 24%. Further, relationship mapping of an additional 178 phenotypic characteristics identified in individuals with FA suggested that approximately 50% of these features could potentially represent novel terms relevant for phenotypic diagnosis of FA, in particular in individuals with *BRCA1*, *BRCA2* or *PALB2* pathogenic variants. The hierarchical relationship of many of the characteristics observed in individuals with FA is unsurprising, and partly reflects the use of multiple HPO terms that provide different levels of detail to describe a “single” given characteristic. It is important to note that ongoing updates to the HPO database can also impact relationship mapping over time. While replication of these potentially novel FA-related terms in independent cohorts may be necessary to formally justify expansion of the ORPHA:84 HPO term list, overall the findings highlight the need to consider “fuzzy matching” ^40^ to improve genetic diagnosis of FA within databases.

Although most individuals in our highly selected cohort presented with one or more recognized FA-related phenotypic features, our findings raise the importance of phenotyping for subtle features to better inform use of recessive disease presentation (or lack thereof) for classification of variants in *BRCA1*, *BRCA2* or *PALB2*. Further, for patients whose clinical features were documented through both previously published reports and prospective data collection, there was a significant difference in the number and characteristics of features reported. It is thus necessary to acknowledge that under-reporting of FA-related features may impact the assignment of the recessive presentation PM3 code for some variants, which is currently weighted based on a combination of chromosome breakage test results, FA physical features, and hematological and toxicity findings. We suggest that use of a standardized data collection instrument, modelled on the one used here for our prospective data collection, would improve the reporting of FA-related features in both general clinical practice and publications.

Another important consideration for PM3 code assignment following the current ClinGen ENIGMA BRCA1 and BRCA2 VCEP specifications is prenatal presentation. Five of the nine individuals with FA diagnosed prenatally were captured by the prospective data collection, perhaps reflecting increasing genetic analysis of suspected abnormalities detected during routine prenatal medical examination. Our data indicates that patients identified prenatally early in gestation, are less likely to develop cancer, and from a practical perspective are less likely to be examined for hematologic or chromosomal abnormalities. This raises the question whether, for the purpose of variant interpretation, prenatal presentation could be considered as a severe clinical entity, and together with other known FA-related physical features, could perhaps be considered as sufficient to assign “recessive clinical features met” for patients biallelic for *BRCA1*, *BRCA2* or *PALB2* variants.

Another component of our study was to assess the possibility that FA-related variants are more likely to be hypomorphic as detected by experimental assays, and/or associated with reduced penetrance in heterozygote individuals. This is an important consideration since the ACMG/AMP classification system overall assumes a Mendelian high-risk disease model, and by inference the PM3 code weight assumes that both variants observed in recessive presentation are associated with dominant disease risk expected for the average pathogenic variant in the relevant gene. Of note, a recent publication reporting a framework to standardize interpretation and reporting of *BRCA1* and *BRCA2* reduced penetrance pathogenic variants^41^ specifically denoted identification of biallelic FA-affected individuals as evidence of a variant associated with reduced penetrance, together with repeated allele observation in compound heterozygote status in individuals with FA, and potential for the allele to partially retain protein function. Our basic categorization of predicted and observed variant type and impact for all FA-related alleles showed that more than 60% of the unique FA-related alleles (and more than 67% of allele observations across families) comprised traditional loss-of-function variant types. Consideration of currently collated functional assay results did not suggest obvious enrichment of hypomorphic missense alleles in individuals with FA. However, genotype-phenotype correlation analysis, building on a previous report demonstrating association of *BRCA2* allele severity score with age at cancer diagnosis in FA individuals,^26^ provided additional insight into allele features that may be used to predict cancer risk. The best-fitting model for predicting age of cancer diagnosis in *BRCA2* FA individuals was achieved by considering potential in-frame mRNA isoform rescue of PTC alleles, replacing “protein” functional results with bioinformatic prediction of protein stability, and annotating both level and predicted/observed in-frame impact of alleles leading to splicing aberrations. Despite the highly significant association achieved by this model, noted examples of between-sib differences in cancer diagnosis raise questions about the clinical utility of the scoring system to predict risk of malignancy in *BRCA2* FA patients at first diagnosis, and invite further study to investigate possible explanations for between-sib differences in presentation e.g. genetic or environmental modifiers. Likewise, further study and much larger cohort sizes will be required to validate these findings, and assess their relevance to predict cancer presentation in *BRCA1* and *PALB2* FA individuals.

Acknowledging such caveats, it is notable that application of an allele severity scoring system to females heterozygous for *BRCA1* or *BRCA2* pathogenic variants revealed evidence for alignment of allele severity score with magnitude of breast cancer risk in the general population. While validation of these findings will be essential, the noted differences in breast cancer risk estimate by *BRCA2* allele severity score could potentially be used to inform differences in risk management for breast cancer. Further, consideration of the association of allele severity score with magnitude of risk for other cancer types, such as ovarian, prostate and pancreatic cancer, will be critical for informing a comprehensive patient management strategy. It is relevant to note that previous segregation analyses comparing breast and ovarian cancer risk associated with *BRCA1* or *BRCA2* pathogenic PTC variants versus missense variants reported evidence that *BRCA1* and *BRCA2* missense alleles overall confer a different breast cancer age-related risk profile, but no obvious differences in ovarian cancer risk profile, compared to *BRCA1* or *BRCA2* PTC variants.^42^ However, in the same study, penetrance analysis limited to 34 families with the *BRCA2* c.7878G>C p.(Trp2626Cys) variant indicated that this FA-related allele (assigned severity score 2 in the final model) was associated with reduced risk of both breast and ovarian cancer compared to the average PTC variant.^42^ Similarly, penetrance analysis focused on the *BRCA1* c.5096G>A p.(Arg1699Gln) variant showed it to be associated with lower breast and ovarian cancer risk compared to the average high-risk pathogenic PTC variant.^35,36^ Further, large-scale detailed penetrance analysis has revealed an atypical cancer risk profile for the FA-related allele *BRCA1* c.5017_5019del p.(His1673del) (allele severity score 2), with heterozygotes demonstrating increased risk of ovarian and uterine cancer, but not breast cancer, in Italian families.^43^ Nevertheless, these findings provide a starting point to explore the concept of allele severity score as a predictor of magnitude of cancer risk associated with *BRCA1* or *BRCA2* alleles considered pathogenic in the recessive versus the dominant state. It will be important to consider alternative or additional experimental data or bioinformatic predictions for allele severity scoring permutations, including exploration of specific functional or mRNA assay methods to distinguish a pattern or range of impact enriched in variants with genetic evidence for reduced (or atypical) cancer risk compared to the average cancer risks associated with *BRCA1* or *BRCA2* pathogenic variants.

Importantly, the current breast cancer risk association findings presented here can be used to at least partly address the question as to whether a FA clinical diagnosis in individuals can provide appropriately weighted evidence for classification of *BRCA1* or *BRCA2* variants under the assumption of a high-risk model. *BRCA1* FA individuals were over-represented for alleles with allele severity score 2, but the magnitude of breast cancer risk for alleles in this category is sufficient to place them in the “high-risk” category. For *BRCA2*, 24% of all FA-related allele combinations included at least one allele with impact severity score 2, a category associated with a 2.6-fold “moderate” level of risk of breast cancer, for which clinical management recommendations are altered compared to classical “high-risk” *BRCA2* variants.^39^ Overall, this would suggest that an FA diagnosis does provide appropriate evidence towards pathogenicity for the majority of *BRCA2* alleles detected in such FA individuals. However, we suggest that that our findings may provide the impetus to introduce allele severity score as an independent factor in future extended variant classification protocols, shifting the current binary pathogenic-benign classification paradigm to better discriminate between clinically actionable “high-risk” pathogenic variants, “reduced/low” penetrance pathogenic alleles, and variants for which clinical action is not justified based on that genetic information alone (including benign variants as well as “risk alleles” falling below clinically actionable thresholds).

Our study was not designed to address whether “lack of FA phenotype” is a reliable negative predictor of *BRCA1*, *BRCA2* or *PALB2* variant pathogenicity, specifically in relation to alleles with reduced (or atypical) penetrance compared to the average pathogenic variant in that gene. However, we note that there is emerging evidence that FA phenotype, assessed against any of the three broad measures currently used in VCEP recommendations, can be absent from individuals compound heterozygote or biallelic for *BRCA1* variants with measurable deleterious impact on function that could confer a clinically significant magnitude of cancer risk. Lack of FA phenotype has been reported for individuals biallelic for the *BRCA1* c.4096+3 A>G allele,^44,45^ a variant that upregulates expression of the *BRCA1* Δ10q in-frame deletion isoform (termed Δ11q using *BRCA1* legacy exon numbering), and has been shown from case-control analysis to be associated with a 3-fold risk of breast cancer and 8-fold risk of ovarian cancer.^46^ As another example, a male without FA phenotype or *BRCA1*- related cancer was identified to be compound heterozygous for a *BRCA1* frameshift variant and an in-frame duplication with impact of transcription activation equivalent to that reported for known “reduced penetrance” variant *BRCA1* c.5096G>A p.(Arg1699Gln).^38^

Together, the results arising from this study have implications for application of *BRCA1*, *BRCA2* and *PALB2* genetic variant information in clinical practice. Overall, our findings have expanded the list of HPO terms to be considered in clinical diagnosis of *BRCA1*, *BRCA2* or *PALB2* related FA, and provided interesting insights into the future *BRCA1* and *BRCA2* genotype-phenotype correlation studies that consider variant type, predicted/observed molecular impact, location of variant and also possible rescue due to in-frame part, single or multi-exon skipping. It will be of interest to conduct large-scale well-powered case-control studies to assess if a similar allele severity scoring approach may provide insight into differences in risk of other (non-breast) cancer types known to be associated with *BRCA1* or *BRCA2* pathogenic variants, and potential differences in cancer risk associated with pathogenic variants in *PALB2* and other cancer predisposition genes.

## Acknowledgements

We are indebted to the generosity of the patients with FA and their families, as well as their treating physicians, for participation in research supporting this study. We thank all the individuals who took part in or enabled the cohort studies informing the breast cancer case-control genetic studies. Additional information relating to acknowledgements and funding sources is provided as **Supplemental Data**.

## Author contributions

Conceptualization, SEJ, KT, JN, MT, AN, EGG, ABS; Methodology, SEJ, MTP, KM, MZ, JN, MT, AN, EGG, ABS; Validation, ABS; Formal analysis, SEJ, ET, KM, MZ, ABS; Investigation, SEJ, ET, MTP, DC, AD, TB, ROR, CPK, RK, LJM, NG, MR, TP, JS, RP, BRV, MG, KNM, KN, SD, MÓF-R, SF, BGdT, MJ, SL, MM, KP, RR, SR, LSB, SELT, KT, ADA, MIC, JAK, MLM, RT, JEW, MW, LD, PJ, TVOH, DE, AS, JN, MT, AN, EGG, ABS; Resources, KM, MZ, TB, ROR, CPK, RK, LJM, NG, MR, TP, JS, RP, BRV, MG, KM, KN, SD, MÓF-R, SF, BGdT, MJ, SL, MM, KP, RR, SR, LSB, SELT, ADA, MIC, JAK, MLM, RT, JFW, MW, NJB, CH, JNW, JH, NR, LRM, MdlH, LW, DE, SS, FC, AS, AN, EGG, ABS; Data Curation, SEJ, ET, MTP, DC, AD, JH, NR, LRM, MdlH, MPGV, LW, SS, FC, ABS; Visualization, SEJ, KM, MZ, DC, AD, LD, JN, MT, EGG, ABS; Supervision, KM, AS, JN, MT, EGG, ABS; Project administration, SEJ, ABS; Funding acquisition, ABS; Writing - Original Draft, SEJ, ABS; Writing - Review & Editing, all authors.

## Web resources

ClinGen Variant Curation Expert Panel (VCEP), https://cspec.genome.network/cspec/ui/svi/

Orphanet rare disease database, https://www.orpha.net/en/disease

Human Phenotype Ontology project, https://hpo.jax.org/.

## Data and code availability

Collated findings for Fanconi anemia individuals are presented in Supplemental Data. Individual HPO terms have not been provided at the patient level for privacy reasons. Such information can be made available from the corresponding author on request, and subject to relevant ethical approvals. BRIDGES data was accessed via the Breast Cancer Association Consortium (BCAC). Individual level data for the BCAC are not publicly available due to ethical review board constraints but are available on request through the BCAC Data Access Co-ordinating Committee. Case-control datasets utilized data from the UK Biobank Resource under application number 102655. Requests for access to UK Biobank data should be made to the UK Biobank Access Management Team. CARRIERS genotype data is available through dbGAP phs002820.v1. Phenotype data are not publicly available due to IRB constraints but are available on request through the CARRIERS Coordinating Committee.

